# ChatGPT as a Digital Pharmacist: A Systematic Review and Meta-Analysis of Drug-Counselling Accuracy

**DOI:** 10.1101/2025.08.10.25333396

**Authors:** Helia Azmakan, Ali Nabipour, Niloufar Ghorabi Tehrani, Niloofar Najari, Pardis Fathi Hafshjani, Alireza Falahati Marvast, Sheida Mani, Negin Asemi Sichani, Samin Fallah Pakdaman, Mobina Shieh, Zeinab Afrandkhalilabad, Arad Shadi, Ramin Shahidi

## Abstract

**Background:** The emergence of Large Language Models (LLMs) like ChatGPT presents significant opportunities for healthcare, yet raises concerns about accuracy, especially in high-risk areas such as medication counseling. A comprehensive evaluation of ChatGPT’s reliability in providing drug information is crucial for its safe integration into clinical practice. This systematic review and meta-analysis aimed to assess the accuracy of drug-counseling information provided by ChatGPT 4.

**Methods:** Following PRISMA, we systematically searched PubMed, Embase, Scopus, and Web of Science on May 9, 2025, for original research evaluating the accuracy of ChatGPT (version 4 or newer) in drug-counseling queries. Included studies compared the AI’s output against standard comparators like pharmacists or drug databases. A random-effects meta-analysis was performed to calculate the pooled proportion of accurate responses, and study quality was assessed using a customized Newcastle-Ottawa Scale (NOS).

**Results:** The search identified 17 eligible studies. Of these, 15 were included in the meta-analysis, which showed a pooled accuracy rate of 86% (95% CI: 0.75–0.95). However, significant heterogeneity was observed across studies (I2=98.5%, p<0.0001). Quality of the studies was a concern, with only four studies (24%) rated as high quality. No evidence of publication bias was found (p=0.91).

**Conclusion:** ChatGPT demonstrates substantial promise in drug counseling, with an 86% accuracy rate that surpasses its performance in other medical domains. However, the high heterogeneity and a non-trivial 14% error rate, coupled with methodological weaknesses in the primary literature, indicate that ChatGPT is not yet ready for autonomous clinical use. Its current role should be as a supplementary tool under the strict supervision of qualified healthcare professionals to ensure patient safety.

## Introduction

The emergence of large language models (LLMs), particularly ChatGPT, has revolutionized the landscape of artificial intelligence applications in healthcare, generating unprecedented interest across medical disciplines and clinical practice settings [1]. Since its public release in 2022, ChatGPT has demonstrated remarkable capabilities in understanding and generating human-like responses to complex medical queries, leading to rapid exploration of its potential applications in clinical decision-making, patient education, and healthcare delivery [2]. The healthcare sector, characterized by its sensitivity to accuracy and stringent regulatory requirements, has witnessed a surge in research investigating the integration of LLMs into various medical domains, from diagnostic support to patient communication [3]. This technological advancement represents a paradigm shift toward general-purpose AI systems that can potentially transform healthcare delivery by providing accessible, immediate, and comprehensive medical information to both healthcare professionals and patients [4]. However, the deployment of LLMs in healthcare settings raises critical questions about accuracy, reliability, and safety, particularly in domains where incorrect information could lead to adverse patient outcomes [5].

Pharmacy practice, as an integral component of the healthcare system, has emerged as a particularly promising domain for AI integration, with artificial intelligence technologies offering significant potential to enhance medication management, clinical decision-making, and patient care [6]. Traditional pharmacy services encompass a wide range of activities including medication reconciliation, drug information provision, adverse drug reaction monitoring, and patient counseling—all of which require extensive pharmaceutical knowledge and clinical expertise [7]. The incorporation of AI technologies, including LLMs like ChatGPT, provides pharmacists and patients with tools that can analyze vast amounts of pharmaceutical data, identify potential drug interactions, assess medication safety profiles, and generate personalized recommendations tailored to individual patient needs [8]. Recent studies have explored ChatGPT’s performance in providing drug dosage information, with some research indicating superior accuracy compared to other AI systems in specific pharmaceutical contexts [9]. However, the complexity of pharmaceutical care, which often involves nuanced clinical considerations, patient-specific factors, and evolving evidence-based guidelines, presents unique challenges for AI-driven drug counseling systems [10].

Despite the promising potential of ChatGPT in pharmaceutical applications, emerging evidence has highlighted significant concerns regarding the accuracy, consistency, and reliability of AI-generated drug information and counseling responses [11]. Research evaluating ChatGPT’s performance in clinical decision-making has revealed substantial limitations, including computational errors, inconsistent responses to identical queries, and the generation of convincing but inaccurate content—a phenomenon known as AI hallucination [12]. Studies specifically examining AI chatbot responses to drug-therapy questions have documented concerning patterns of inaccuracy, particularly in complex clinical scenarios requiring detailed explanations or evidence-based recommendations. The LLMs’ ability to produce harmful or misleading medical information, combined with their propensity to generate non-existent citations and references, poses significant risks in pharmaceutical counseling contexts where accuracy is paramount for patient safety [13]. Furthermore, systematic reviews of ChatGPT’s applications in healthcare have consistently emphasized the need for rigorous validation, human oversight, and ethical guidelines to ensure safe implementation in clinical practice [14].

Given the rapid adoption of ChatGPT and similar LLMs in healthcare settings, coupled with the critical importance of accurate drug information in pharmaceutical care, there is an urgent need for systematic evaluation of these technologies’ performance in drug counseling applications. While individual studies have examined specific aspects of ChatGPT’s pharmaceutical knowledge, a comprehensive synthesis of evidence regarding its accuracy in drug counseling contexts remains lacking. This systematic review and meta-analysis aims to address this knowledge gap by systematically evaluating the accuracy of drug information provided by ChatGPT compared to standard pharmacist counseling and established drug information resources.

## Methods

This systematic review and meta-analysis was conducted and reported in accordance with the Preferred Reporting Items for Systematic Reviews and Meta-Analyses (PRISMA) 2020 guidelines [15]. The review protocol was registered with PROSPERO (ID: CRD420251049637).

### Search Strategy

A systematic search was conducted on May 9, 2025, across four electronic databases: PubMed, Embase, Scopus, and Web of Science. The search strategy combined terms for “ChatGPT” with keywords related to pharmaceuticals and medication. The exact search strategies used were as follows: (“Chat-GPT” OR “Chat GPT” OR “ChatGPT*”) AND (“drug*” OR “medication*” OR “pharmac*” OR “prescription*”))

### Study Selection

All records were imported into a Rayyan web application [16] for manual deduplication. Two reviewers independently screened the titles and abstracts for eligibility. The full texts of potentially relevant articles were then retrieved and assessed by the same two reviewers against the inclusion criteria. Any disagreements were resolved through discussion or, if necessary, by consulting a senior reviewer (RS).

This review included original, English-language research studies that evaluated the accuracy of drug information provided by ChatGPT (version 4 or newer). To be eligible, studies were required to focus on queries from patients or healthcare professionals and compare the AI’s output against a standard comparator, such as pharmacist counseling, drug databases, or clinical guidelines. We excluded studies that were not original research (e.g., editorials, reviews, preprints) or did not report on accuracy as a primary outcome. Furthermore, to maintain a focused scope on medication counseling, studies centered on other tasks—such as prescribing, deprescribing, answering exam questions, or identifying medications from images—were excluded, as were those that assessed custom-built language models or evaluated information on traditional or herbal medicine.

### Data Extraction and Quality Assessment

A standardized data extraction form was used to collect key information from each included study, including publication details, study design, country, sample size, comparator type, and prompting strategy, and limitations. For the meta-analysis, we also extracted the total number of queries and the number of queries answered correctly. The methodological quality of the included studies was independently assessed by two reviewers using a customized Newcastle-Ottawa Scale (NOS) [17]. This tool was adapted to evaluate the risk of bias by assessing three domains: 1) Representativeness and Study Design, 2) Comparability and Bias Control, and 3) Outcome and Assessment. Based on the final score, studies were categorized as having a High, Moderate, or Low quality.

### Statistical Analysis

For the quantitative analysis, a meta-analysis of proportions was performed using R (version 4.5) with the ‘meta’ package. Due to possible heterogeneity in this research context, a random-effects model was employed to calculate the pooled proportion of accurate responses and its corresponding 95% confidence interval (CI). Heterogeneity was quantified using the I^2^ statistic, with thresholds of 25%, 50%, and 75% considered to represent low, moderate, and high levels of heterogeneity, respectively. Publication bias was assessed via visual inspection of a funnel plot and formally with Egger’s linear regression test, where a p-value < 0.10 was considered indicative of significant asymmetry.

## Results

### Study Selection

The initial search across four databases (Scopus, Web of Science, PubMed, and Embase) identified a total of 1,453 records. After the removal of 773 duplicate entries, 680 unique records proceeded to the screening stage. During the screening of titles and abstracts, 595 records were excluded. Of these, 497 were rejected based on title screening and an additional 98 were removed following abstract screening. This process resulted in 85 articles being selected for full-text eligibility assessment. Upon full-text review, 68 articles were further excluded for specific reasons. The most common reasons for exclusion were being a non-original study (e.g., review, editorial) (n=35) or assessing ChatGPT version 3.5 (n=24). Other reasons for exclusion included the full text being unavailable (n=3), the study assessing multiple-choice questions (MCQ) (n=3), the article not being in English (n=2), and one study focusing on prescribing drugs (n=1). After this comprehensive selection process, a final total of 17 studies met the inclusion criteria and were included in this review. **(Figure 1)**

**Figure 1.**
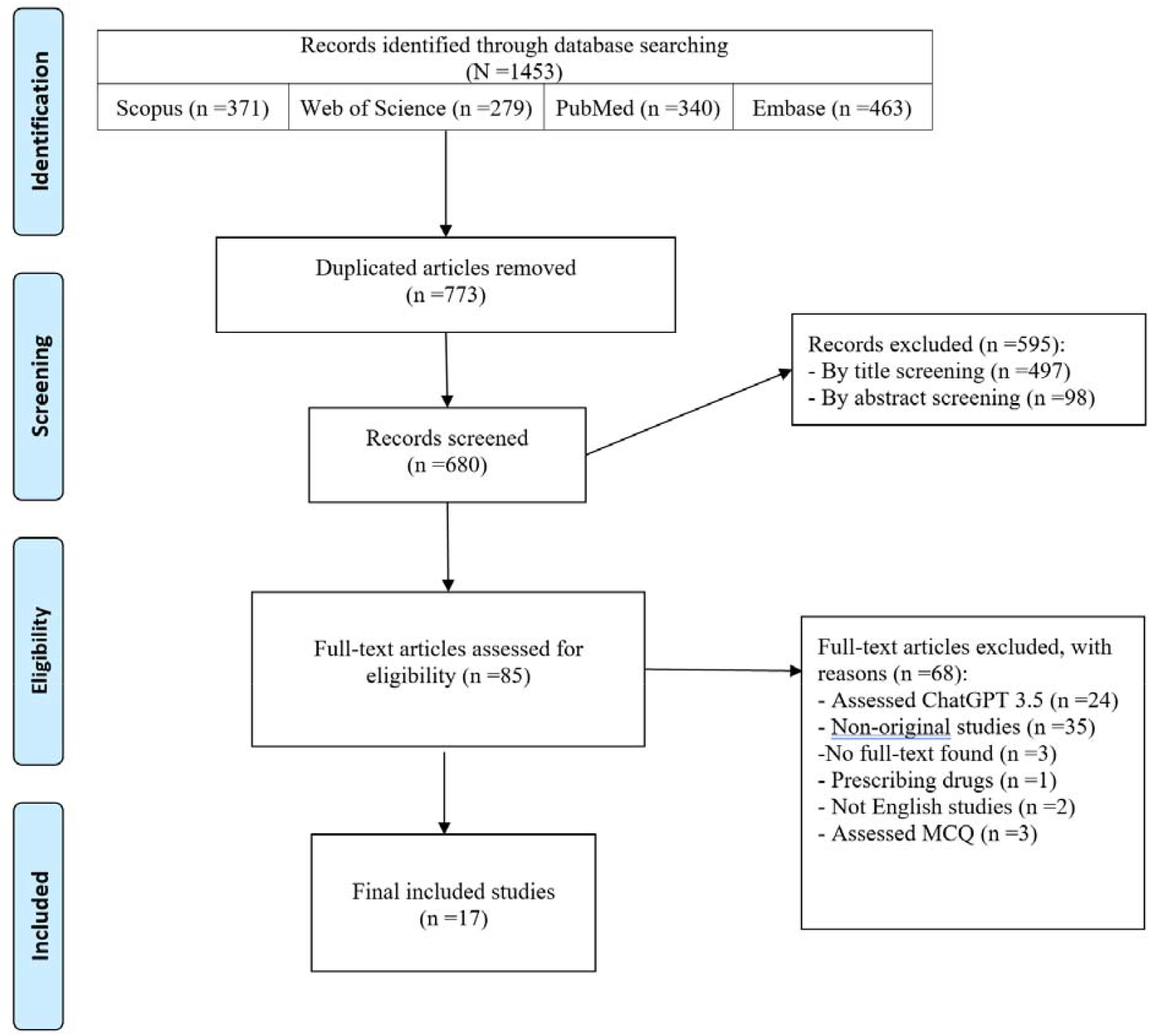
PRISMA 2020 flow diagram illustrating the study selection process.

### Characteristics of Included Studies

The characteristics of the 17 studies included in this systematic review are detailed in **Table 1**. The studies were recent, with publication years ranging from 2023 to 2025, highlighting the contemporary interest in this topic. The research was geographically diverse, originating from 12 different countries across North America, Europe, Asia, and the Middle East, with the highest number of studies coming from the USA (n=4).

**Table 1.**
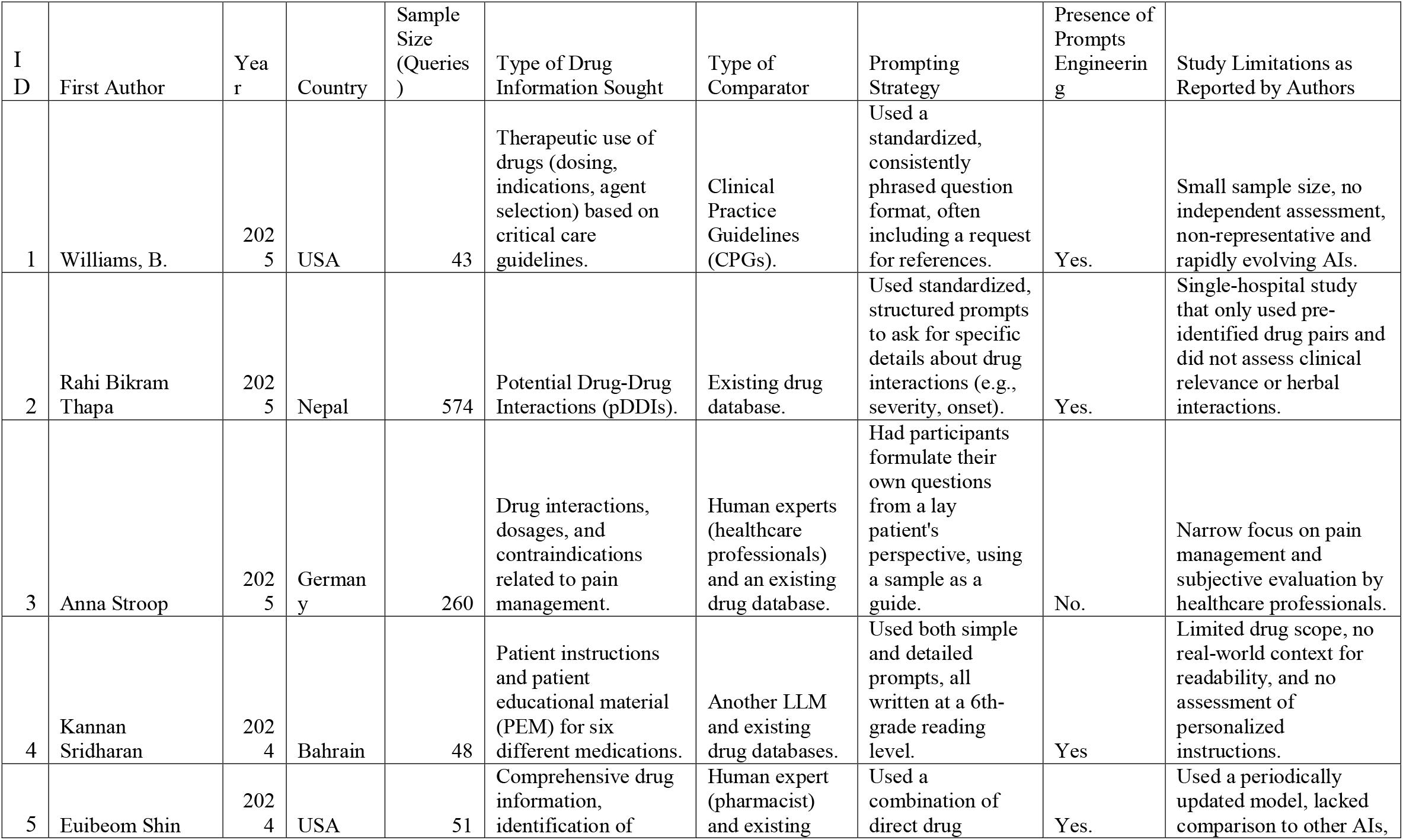
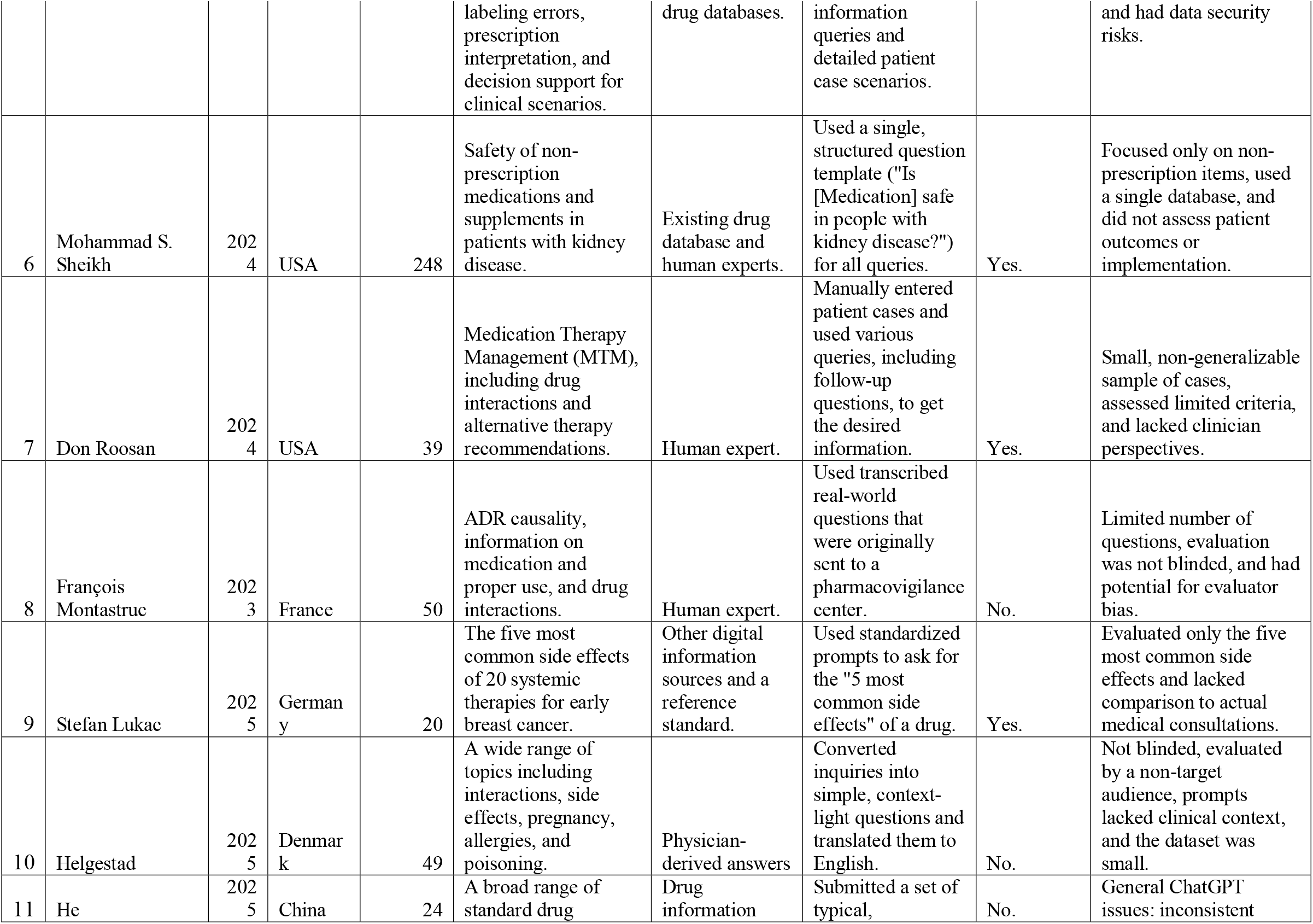
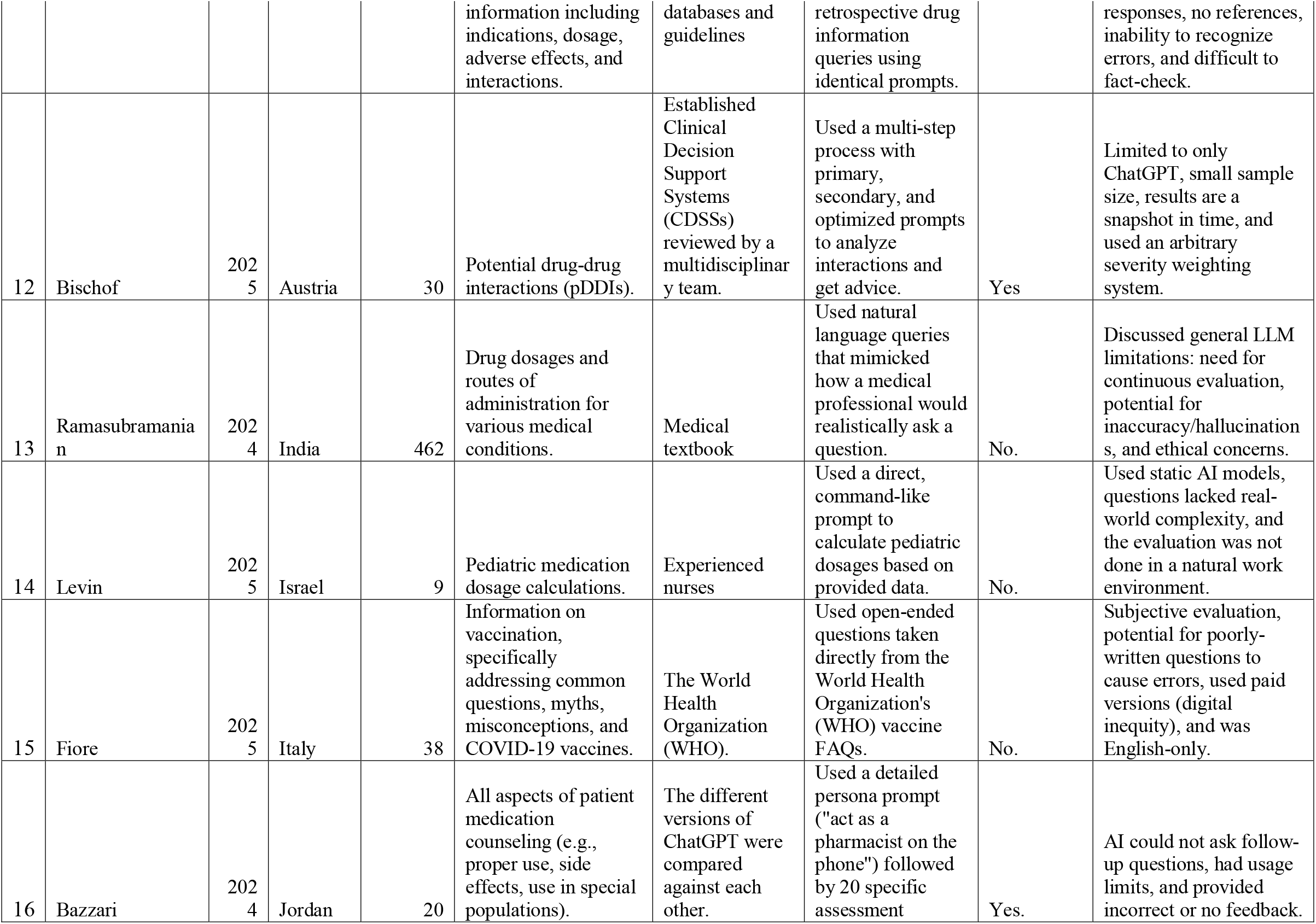
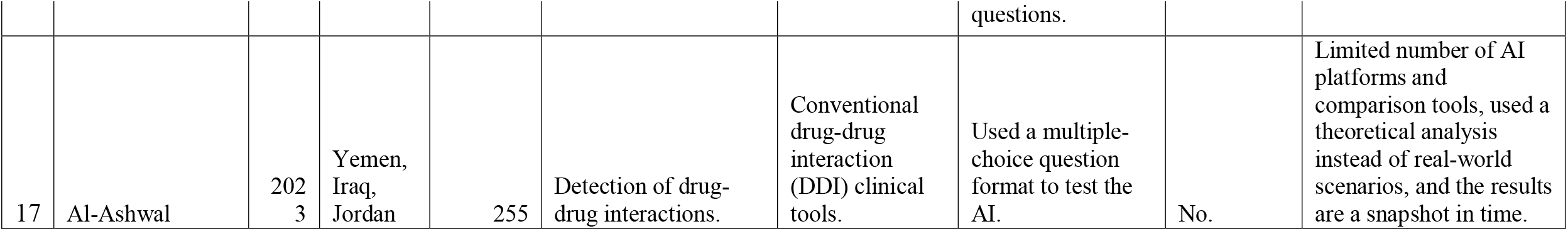
Characteristics of the 17 studies included in the systematic review.

The number of queries used to evaluate the AI models varied substantially across the studies, ranging from as few as 9 to as many as 574. The research questions explored a wide spectrum of drug information needs. The most frequently investigated topics included: 1. Potential Drug-Drug Interactions (pDDIs): Investigated in five studies (Thapa, Stroop, Montastruc, Bischof, Al-Ashwal). 2. Drug Dosage and Administration: Assessed in five studies (Williams, Stroop, Ramasubramanian, Levin, He). 3. General Drug Information and Patient Counseling: Covered in four studies (Sridharan, Shin, Bazzari, He).

Other studies focused on more specialized areas such as therapeutic use in critical care (Williams), safety in kidney disease (Sheikh), Medication Therapy Management (MTM) (Roosan), adverse drug reactions (Montastruc, Lukac), and vaccination information (Fiore).

To benchmark the performance of the AI, studies utilized a variety of comparators. The most common were established drug information databases or Clinical Decision Support Systems (CDSSs) (n=8) and human experts such as pharmacists, physicians, or nurses (n=7). Other benchmarks included official guidelines, medical textbooks, and other large language models.

A significant methodological difference was the approach to prompting. Ten of the 17 studies employed prompt engineering, using structured, standardized, or multi-step query formats to optimize the AI’s output. For example, some used persona prompts (Bazzari) or detailed question templates (Sheikh, Lukac). In contrast, seven studies used a more organic approach without explicit prompt engineering, often using real-world questions from patients and clinicians (Montastruc, Helgestad) or allowing participants to formulate their own queries (Stroop).

The authors of the included articles frequently acknowledged several key limitations. The most common were small sample sizes and a narrow focus on specific drugs or conditions, which may limit the generalizability of the findings. Another widely reported issue was that the results represent a “snapshot in time” of a rapidly evolving AI technology. Other recurring limitations included the lack of real-world clinical context, the potential for evaluator bias in non-blinded assessments, and inherent LLM weaknesses such as the potential for generating inaccurate information (“hallucinations”) and the inconsistency of responses.

### Quality Assessment

The methodological quality of the 17 included studies was evaluated using a customized Newcastle-Ottawa Scale (NOS) adapted for cross-sectional studies, with a maximum possible score of 10 stars. Based on their total scores, studies were categorized as being of high, moderate, or low quality. Details of this assessment provided in **Table 2**.

**Table 2.**
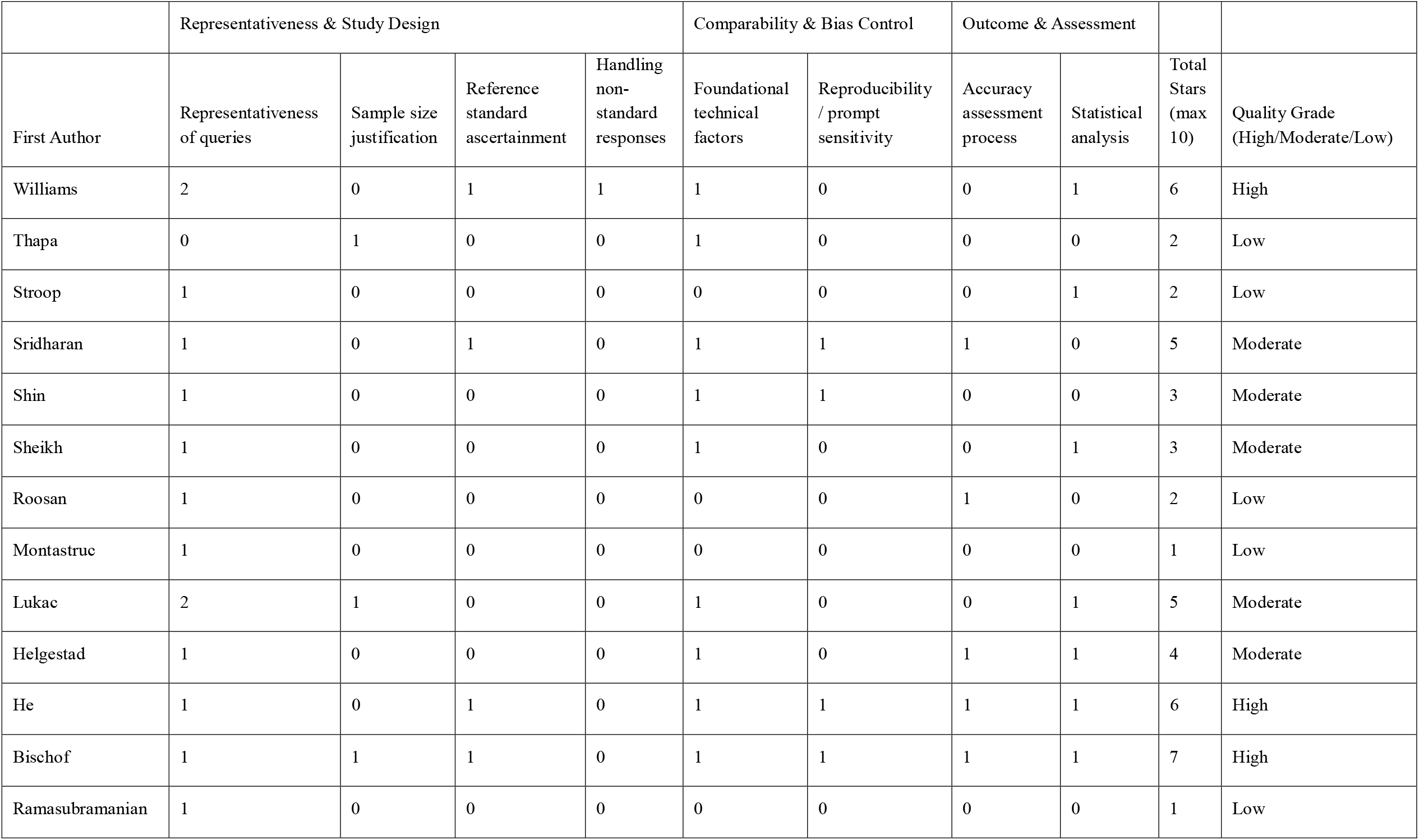
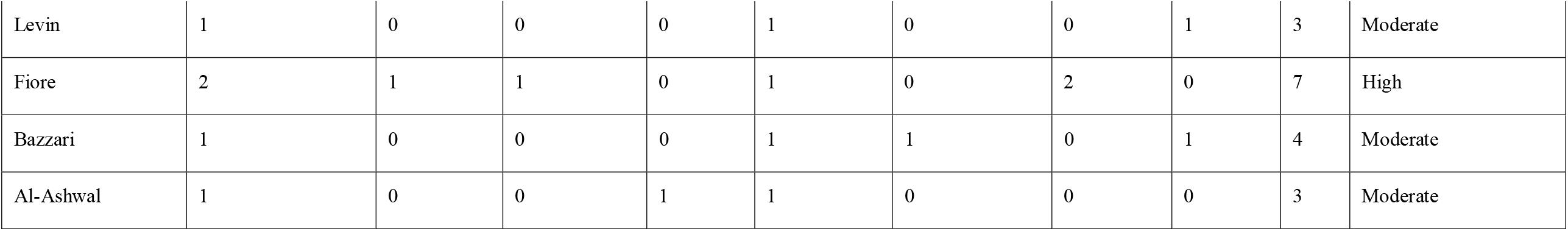
Methodological quality assessment of included studies using the customized Newcastle-Ottawa Scale (NOS).

Overall, four studies (24%) were rated as High quality, with scores ranging from 6 to 7 stars. The majority of the articles, eight studies (47%), were deemed to be of Moderate quality, with scores between 3 and 5 stars. The remaining five studies (29%) were classified as Low quality, scoring 2 or fewer stars. A detailed analysis of the scoring criteria revealed several common patterns across the included research. A significant and common weakness was the lack of sample size justification; only four studies (24%) received a point for this criterion. Other frequently missed criteria included:

- Handling non-standard responses: Only two studies (12%) had a clear protocol for managing incomplete, irrelevant, or non-standard answers from the AI.
- Reproducibility/Prompt sensitivity: Only five studies (29%) assessed whether minor changes to the prompts would alter the AI’s responses.
- Accuracy assessment and statistical analysis: Fewer than half of the studies employed a robust, blinded, or independent accuracy assessment process (n=8) or conducted formal statistical analysis (n=8).

### Meta-Analysis results

A meta-analysis was performed on 15 studies, to calculate the pooled proportion of accurate or appropriate responses. The overall pooled proportion of accurate responses was 0.86 (86%), with a 95% confidence interval (CI) of 0.75 to 0.95. The analysis revealed a high degree of heterogeneity between the studies, with an I2 statistic of 98.5% (p<0.0001). **(Figure 2)** This indicates that while the average performance is high, the results varied considerably across the different study conditions. Publication bias was investigated through visual inspection of a funnel plot and formally with Egger’s linear regression test. The funnel plot showed that the studies were distributed relatively symmetrically around the mean effect estimate, suggesting a low likelihood of publication bias. This visual finding was supported by the statistical test, which found no evidence of significant funnel plot asymmetry (t=−0.12, p=0.91). **(Figure 3)** These results indicate that the pooled estimate is unlikely to be substantially affected by publication bias.

**Figure 2.**
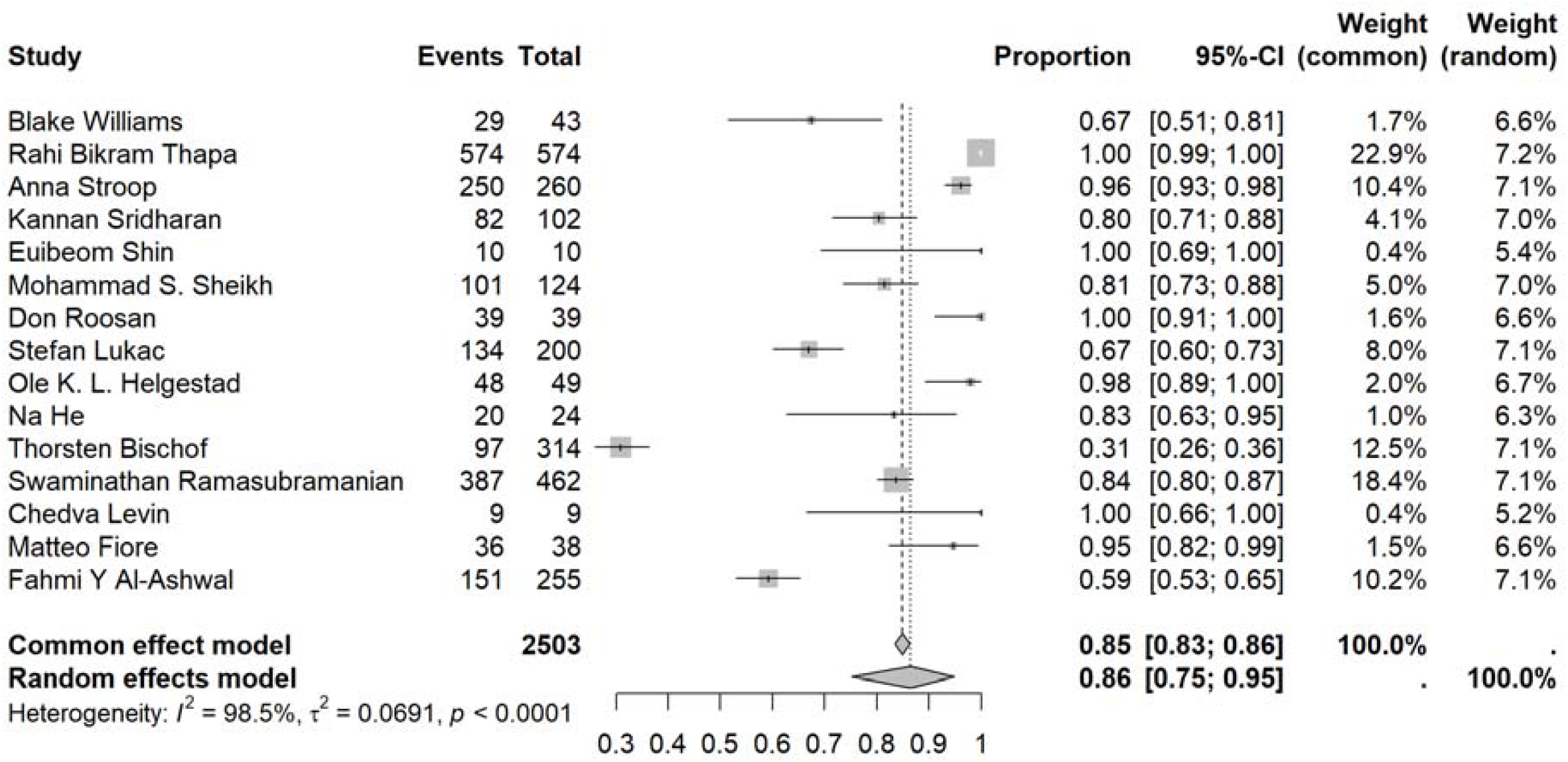
Forest plot of the meta-analysis on the proportion of accurate responses.

**Figure 3.**
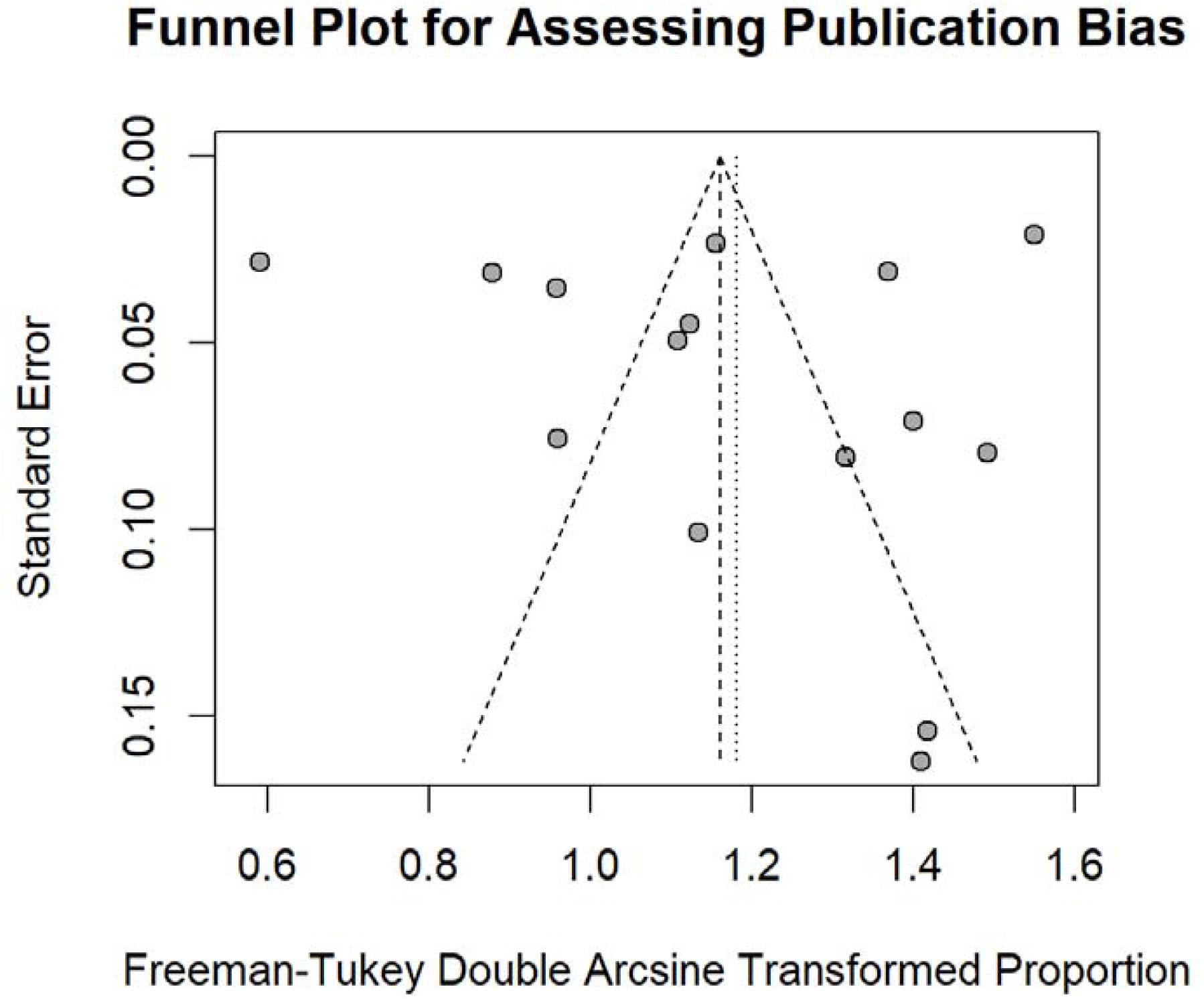
Funnel plot for the assessment of publication bias.

## Discussion

This systematic review and meta-analysis provides the first comprehensive assessment of ChatGPT’s performance as a digital pharmacist, analyzing 17 studies with 2,503 drug-related queries. Our meta-analysis revealed an overall accuracy rate of 86% (95% CI: 0.75-0.95), indicating that ChatGPT demonstrates substantial competency in pharmaceutical counseling applications. The included studies, published between 2023-2025, examined diverse domains including potential drug-drug interactions (five studies), dosage and administration guidance (five studies), and general drug information counseling (four studies). Despite the encouraging accuracy rate, we observed high heterogeneity (I^2^=98.5%, p<0.0001), suggesting considerable variation in performance across different pharmaceutical scenarios and study methodologies.

Quality assessment using our adapted Newcastle-Ottawa Scale revealed significant methodological limitations, with only four studies (24%) achieving high quality ratings. The most common deficiencies included inadequate sample size justification (76% of studies), insufficient protocols for handling non-standard AI responses (88% of studies), and limited assessment of prompt reproducibility (71% of studies). Notably, ten studies employed prompt engineering techniques, which appeared to enhance ChatGPT’s performance compared to naturalistic query approaches. Our assessment found no evidence of publication bias (Egger’s test: t=-0.12, p=0.91), strengthening confidence in the pooled accuracy estimate.

The observed 86% accuracy rate can be attributed to several factors that distinguish pharmaceutical information from other medical domains. Unlike clinical diagnosis or treatment planning, drug counseling often involves retrieval and synthesis of well-established, standardized information from authoritative pharmaceutical databases and formularies [6]. This structured nature of pharmaceutical knowledge aligns favorably with large language models’ pattern recognition and information synthesis capabilities, as demonstrated in previous AI applications in pharmacy practice [7].

The substantial impact of prompt engineering on performance suggests that the high accuracy rate may reflect optimized query formulation rather than robust real-world performance. Studies using persona prompts or structured questioning approaches consistently reported higher accuracy than those employing conversational queries [12]. This finding raises important questions about ChatGPT’s reliability when users formulate queries spontaneously without structured prompting, as would occur in typical clinical scenarios.

The exclusion of ChatGPT-3.5 studies from our analysis likely contributed to the favorable accuracy rates observed. Research comparing different ChatGPT versions has consistently demonstrated superior performance of GPT-4, with accuracy improvements of 20-30 percentage points over earlier versions in medical applications [18, 19]. The high heterogeneity observed across studies reflects the genuine complexity of pharmaceutical counseling scenarios, ranging from simple drug identification to complex interaction assessments requiring clinical judgment [20].

Our findings of 86% accuracy in drug counseling substantially exceed ChatGPT’s documented performance in other healthcare domains. Wei et al.’s comprehensive meta-analysis of ChatGPT in general medical applications reported only 56% overall accuracy (95% CI: 51-60%), with internal medicine achieving 63% and surgery 49% [21]. This 30 percentage point advantage suggests that pharmaceutical applications represent a particularly suitable domain for AI implementation in healthcare.

Similarly, Liu et al.’s analysis of ChatGPT performance in medical licensing examinations found GPT-4 achieved 81% accuracy compared to GPT-3.5’s 58% [19]. While our 86% accuracy approaches the upper range of medical examination performance, the comparison highlights the importance of version-specific evaluation and the superior capabilities of advanced language models. The consistently superior performance in pharmaceutical applications compared to general medical domains (86% vs 56%) suggests that drug counseling may represent an optimal entry point for healthcare AI integration. This advantage likely reflects the availability of high-quality pharmaceutical training data, standardized reference materials, and the factual nature of much drug information compared to the complex reasoning required for clinical diagnosis or treatment planning.

### Limitations

Several important limitations affect the interpretation and generalizability of our findings. The variable study quality, with 76% of studies lacking adequate methodological rigor, raises concerns about the reliability of individual accuracy estimates and our pooled results. The rapid evolution of ChatGPT technology means our findings represent temporal snapshots that may not reflect future capabilities, particularly given the continuous model updates and improvements occurring during our study period. The controlled experimental conditions under which most studies were conducted may not adequately reflect real-world implementation challenges, where queries are formulated spontaneously, evaluation resources are limited, and integration with existing healthcare systems is required. The absence of patient-centered outcome measures represents a critical limitation for assessing clinical relevance. While accuracy metrics provide important information about response correctness, they do not capture patient comprehension, satisfaction, adherence to recommendations, or ultimate clinical outcomes. The diversity of evaluation methodologies and comparison standards across studies further complicates interpretation, with different benchmarks potentially yielding varying accuracy assessments for identical AI responses.

## Conclusion

ChatGPT shows high accuracy (86%) in drug counseling, suggesting its potential as a digital pharmacist tool. However, a 14% error rate and extreme variability mean it’s not ready for independent clinical use. Its best application is as a supplementary tool with robust human oversight, not for autonomous decisions. The structured nature of drug information makes it an ideal domain for AI integration. Future work requires standardized evaluation and regulatory guidelines to ensure safety. A cautious approach using hybrid human-AI systems is the most promising path forward.

## Data Availability

All data produced in the present study are available upon reasonable request to the authors

## Notes

### Competing Interest Statement

The authors have declared no competing interest.

### Funding Statement

This study did not receive any funding

